# Prevalence and Determinants of Diabetes among Middle-Aged and Older Adults in India: A Cross-Sectional Analysis of LASI

**DOI:** 10.64898/2026.10.31.26366972

**Authors:** Rajeev Kumar, Suraj Prakash, Simran Kumari

## Abstract

**Background:** Diabetes is an increasing public health challenge in India, particularly among middle-aged and older adults. Understanding its prevalence and associated risk factors is essential for informing prevention and management strategies.

**Objective:** This study aimed to estimate the prevalence of diabetes and identify demographic, socioeconomic, behavioural, and health-related factors associated with diabetes among adults aged 45 years and above in India.

**Methods:** A cross-sectional analysis was conducted using nationally representative data from the Longitudinal Ageing Study in India (LASI) Wave-1 (2017–18), comprising 72,055 individuals aged 45 years and above. Diabetes status was based on respondents’ self-reported diagnosis by a health professional. Descriptive statistics were used to estimate diabetes prevalence, and multivariable binary logistic regression was performed to examine associations between diabetes and selected covariates, including age group, sex, place of residence, caste, religion, physical activity, dietary practices, heart disease, and heart attack risk.

**Results:** Diabetes prevalence was higher among older adults, urban residents, females, and individuals belonging to the Other Backward Classes (OBC). After adjustment for potential confounders, age, urban residence, caste, religion, heart disease, and heart attack risk were significantly associated with diabetes, whereas sex was not statistically significant after adjustment. Individuals aged 55–64 years and those residing in urban areas exhibited significantly higher odds of diabetes compared with their respective reference groups.

**Conclusions:** Diabetes is highly prevalent among middle-aged and older adults in India and is associated with several demographic, socioeconomic, and health-related factors. These findings highlight population groups that may benefit from targeted prevention, screening, and management strategies. Further longitudinal studies are warranted to clarify temporal relationships and causal pathways.

## 1. Introduction

Diabetes mellitus is one of the fastest-growing non-communicable diseases worldwide and represents a major public health challenge because of its increasing prevalence, long-term complications, and substantial economic burden. The International Diabetes Federation estimates that more than 537 million adults were living with diabetes in 2021, with this number projected to increase markedly over the coming decades. Low- and middle-income countries account for most of this increase, reflecting rapid population ageing, urbanisation, and lifestyle transitions (Bjork & Stefan (2003), Anjana et al., 2011).

India has one of the largest populations of people living with diabetes. Rapid demographic change, urbanisation, reduced physical activity, dietary transitions, and increasing obesity have contributed to a substantial rise in diabetes prevalence over recent decades (Hu F. B., 2011). Diabetes is associated with serious complications, including cardiovascular disease, chronic kidney disease, visual impairment, and premature mortality, thereby imposing considerable pressure on individuals, households, and the healthcare system (Dal canto E. et al.,2019). Previous studies have consistently shown that diabetes prevalence varies according to age, sex, place of residence, socioeconomic status, and lifestyle behaviours, although the magnitude of these associations differs across populations (Bjork & Stefan, 2003; Anjana R. M., et al., 2017; Deshpande et al., 2008).

Several nationally representative studies, including the ICMR–INDIAB study and the National Non-Communicable Disease Monitoring Survey, have documented the growing burden of diabetes in India (Gyawali B. et al., 2018). However, relatively few studies have comprehensively examined the combined influence of demographic, socioeconomic, behavioural, and health-related characteristics among middle-aged and older adults using data from the Longitudinal Ageing Study in India (LASI). Given India’s rapidly ageing population, identifying population groups with a higher prevalence of diabetes is important for planning prevention and disease management strategies (Wanga. Et al., 2018 and Wong E. et al., 2013). The Longitudinal Ageing Study in India (LASI) provides nationally representative information on the health, social, and economic conditions of adults aged 45 years and above Hunt, D. et al., 2021). Its extensive information on chronic diseases, behavioural characteristics, and socioeconomic conditions enables a comprehensive assessment of factors associated with diabetes among middle-aged and older adults (Khan M. A.B et al., 2020).

Therefore, the present study aims to estimate the prevalence of diabetes among adults aged 45 years and above in India and to examine its association with demographic (age, sex, place of residence, caste, and religion), behavioural (physical activity and dietary practices), and health-related factors (heart disease and heart attack history) using nationally representative LASI Wave-1 (2017–18) data through multivariable binary logistic regression analysis. In this paper, we divide it into multiple sections and subsections which is as section 1 includes the introduction and subsections of section has included related works, In section 2 includes Methodology and its subsections includes the Data source, analytical techniques and Data table of prevalence of Diabetes which is from LASI, In section 3, includes Data representation by the help of chart, In section 4 includes Results and Discussion by the help of Binomial Logistic Regression prevalence significance table and finally in section 5 include Concluding Remarks of this study and also in lastly included reference which is used in this study.

## 2. Related Works

Some key areas of research and Literature on **Condition of Diabetes in India** include: - Diabetes is the most common non-communicable disease globally. The estimated number of adults with diabetes in 2007 was 246 million of these, 80% live in developing countries, the largest numbers on the Indian subcontinent and in China. Approximately 85-95% of all cases of diabetes are type 2 diabetes and the worldwide explosion of this disorder is a major health care burden. It is estimated that nearly 380 million adults worldwide will have diabetes by 2025. India has 41 million diabetics, and this number is expected to increase to 70 million by 2025.The increased number of diabetics in India is likely to be due to a significant increase in the incidence of type 2 diabetes, caused by unprecedented rates of urbanization, which results in environmental and lifestyle changes (Krishnan.A. et al., 2021). According to World Health Organization (WHO) estimates, the urban population in developing regions will increase from 1.9 billion in 2000 to 3.9 billion in 2030. It is estimated that, by 2030, nearly 46% of India’s population will be living in urban areas. Chronic diseases, such as diabetes and cardiovascular disease (CVD), pose a primary challenge for the health care system. India is the second most populous country, with considerable diversity in caste, religion, habitat, and socioeconomic status, lifestyle, and food habits. Although several infectious and parasitic diseases have been controlled successfully in India, non-communicable diseases are becoming increasingly common, resulting in an enormous burden on the health care system. Significant economic improvements have occurred in India, but the large population creates difficulties for the effective reduction of poverty, malnutrition, and the provision of health care to all. India, China, and Pakistan contribute 21%, 12%, and 5%, respectively, to the annual increase in the global population (Ramachandran Ambady, 2012). India, a country experiencing rapid socioeconomic progress and urbanization, carries a considerable share of the global diabetes burden. Studies in different parts of India have demonstrated an escalating prevalence of diabetes not only in urban populations, but also in rural populations because of the urbanization of lifestyle parameters (Tandon, N. et al., 2018). The prevalence of prediabetes is also high ( Kumar,A, et al.,2024). Recent studies have shown a rapid conversion of impaired glucose tolerance to diabetes in the southern states of India, where the prevalence of diabetes among adults has reached approximately 20% in urban populations and approximately 10% in rural populations (Kumar, A. et al., 2024 and Maiti, S. et al., 2023). Because of the considerable disparity in the availability and affordability of diabetes care, as well as low awareness of the disease, the glycaemic outcome in treated patients is far from ideal. Lower age at onset and poor glycaemic control are likely to increase the risk of vascular complications (Nanditha Arun, 2012). The economic burden of treating diabetes and its complications is considerable. It is appropriate that the Indian Government has initiated a national program for the management and prevention of diabetes and related metabolic disorders. Lifestyle modification is an effective tool for the primary prevention of diabetes in Asian Indians (Ramachandran Ambady, 2012 and Lee, j. et al., 2021). According to World Health Organization (WHO) the number of people with diabetes rose from 108 million in 1980 to 422 million in 2024. Prevalence has been rising more rapidly in Low- and middle-Income countries than in High Income countries. Diabetes is Major Cause of Blindness, kidney failure, Heart attacks, stroke and lower limb amputation. A healthy diet, regular physical Activity, maintaining a Normal Body weight and avoiding tobacco use are ways to prevent or delay the onset of Type2 Diabetes It is recommended that a person with Diabetes actively engage themselves in self- management, education, and treatment planning along with an individualised diet plan ( Mohan, V. et al., 2007). With India being the Diabetes capital of the world, it is high time that people with Diabetes follow a diabetic diet chart, as it significantly helps maintain blood glucose levels and alleviate Diabetes symptoms (Lin, X. et al., 2020). A good diet plan for diabetics includes more non-starchy vegetables, such as broccoli, spinach and green beans. Fewer added sugars and refined grains white bread, rice and pasta with less than two grams of fibre external icon per serving (Rahman, M.S et al., 2011). Carbohydrates raise your blood sugar levels and how fast they do it depends on what food you eat. For instance, drinking fruit juice raises blood sugar faster than eating whole fruit. Also, eating carbs with foods that have protein, fat or fibre slows down how quickly your blood sugar rises. This intake needs to be monitored and planned (Stokes, A et al., 2017). Regular and balanced meals help to avoid high or low blood sugar levels. You must eat the same amount of carbs at each meal as this helps the diabetes diet plan (Liu, X. et al., 2016). The plate method and counting carbs are two common tools that can make planning meals simple. Also, meal planning may vary with the kind of diabetes you are diagnosed with (Murugans, N. et al., 2007). As the burden of diabetes on total health care spending is likely to increase and, potentially, will have important consequences on the sustainability of health care financing, this study presents a critical review of the literature on cost of illness of diabetes and its complications in India and makes recommendations on areas requiring further attention and research (Maiti, S. et al., 2023). Diabetes is a severe chronic disease that arises when insulin generation is insufficient, or the generated insulin cannot be used in the body, resulting a long-term metabolic disorder. Diabetes affects an estimated 537 million adults worldwide between the ages of 20 and 79 (10.5% of all adults in this age range). By 2030, 643 million people will have diabetes globally, increasing to 783 million by 2045. According to the IDF 10th edition, the incidence of diabetes has been rising in Southeast Asia (SEA) nations for at least 20 years, and current estimates have outperformed all previous forecasts (Kumar Arvind, 2024).

According to Mishra, A. et al.(2019) and Misra,A & Khurana L(2008) estimated prevalence of diabetes in adults (20 to 79 years) has more than tripled since the first edition in 2000, rising from an estimated 151 million (4.6% of the world’s population at the time) to 537.5 million (10.5%) of the world’s population today. Beside of this according to Kumar Arvind, (2024) prevalence rate will be higher than 12.8% by 2045. In addition, this study indicates that the incidence of diabetes in the world, Southeast Asia, and India was 10.5%, 8.8%, and 9.6%, respectively, throughout 2021 and will rise to 12.5%, 11.5%, and 10.9%, respectively by 2024.

## 3. Methodology

### 3.1 Study Design and Data Source

This study is a **cross-sectional secondary analysis** of data obtained from the **Longitudinal Ageing Study in India (LASI) Wave-1 (2017–18)**. Although LASI is a longitudinal cohort study, the present analysis uses only the baseline survey; therefore, all findings are based on cross-sectional data. LASI is a nationally representative survey conducted by the International Institute for Population Sciences (IIPS), Mumbai, in collaboration with the Ministry of Health and Family Welfare, Government of India, and several international partners. The survey collected detailed information on demographic characteristics, socioeconomic conditions, physical and mental health, healthcare utilisation, chronic diseases, and health-related behaviours among adults aged **45 years and above** and their spouses across all states and union territories of India. The present study included **72,055 respondents aged 45 years and above** with complete information on diabetes status and selected explanatory variables. Individuals with missing information on the outcome or key covariates were excluded from the multivariable analysis.

### 3.2 Outcome Variable

The outcome variable was **self-reported diabetes mellitus**. Respondents were classified as having diabetes if they answered **“Yes”** to the LASI question asking whether a doctor or other health professional had ever diagnosed them with diabetes or high blood sugar. The outcome variable was coded as:

- 1 = Diabetes
- 0 = No diabetes

### 3.3 Explanatory Variables

Based on previous epidemiological studies and the availability of variables in LASI, the following explanatory variables were included in the analysis:

- Age group (<45, 45–54, 55–64, 65–74, and ≥75 years)
- Sex (Male/Female)
- Place of residence (Rural/Urban)
- Caste (SC, ST, OBC, Others)
- Religion (Hindu, Muslim, Christian, Others)
- Physical activity (Daily, Weekly, Never)
- Special diet (Yes/No)
- Heart disease (Yes/No)
- Heart attack history (Yes/No)

If respondents younger than 45 years were retained because they were spouses of age-eligible participants, this has been explicitly acknowledged. Otherwise, this category should be excluded from all analyses to maintain consistency with the stated study population.

### 3.4 Statistical Analysis

Descriptive statistics were used to summarize the characteristics of the study population and estimate the prevalence of diabetes across demographic, socioeconomic, behavioural, and health-related characteristics. Pearson’s chi-square test was used to assess bivariate associations between diabetes status and categorical explanatory variables.

Multivariable binary logistic regression analysis was performed to identify factors independently associated with diabetes. Variables included in the adjusted model were selected based on previous epidemiological evidence and their established relevance to diabetes. Adjusted odds ratios (AORs) and 95% confidence intervals (CIs) were estimated. Statistical significance was assessed at a two-sided *p* < 0.05.

All analyses were performed using IBM SPSS Statistics (Version XX). Data management and figure preparation were conducted using Microsoft Excel.

### 3.5 Ethical Considerations

This study used publicly available, de-identified LASI Wave-1 data. Ethical approval for data collection was obtained by the LASI investigators from the Institutional Review Board of the International Institute for Population Sciences (IIPS), Mumbai, and informed consent was obtained from all participants before data collection. Since the present study involved secondary analysis of anonymized data, no additional ethical approval was required.

**Table 3.1** presents the baseline characteristics of the **72,055 adults aged 45 years and above** included in the analysis. Overall, **11.6%** of participants reported having been diagnosed with diabetes. Diabetes prevalence increased with age and was highest among participants aged **65–74 years**, followed by those aged **55–64 years**. The prevalence was higher among **women**, **urban residents**, participants belonging to the **Other Backward Class (OBC)** category, and **Christians**. Significant differences in diabetes prevalence were also observed according to physical activity, special diet, heart disease, and heart attack history (**all p < 0.05**).

**Table 3.1.**
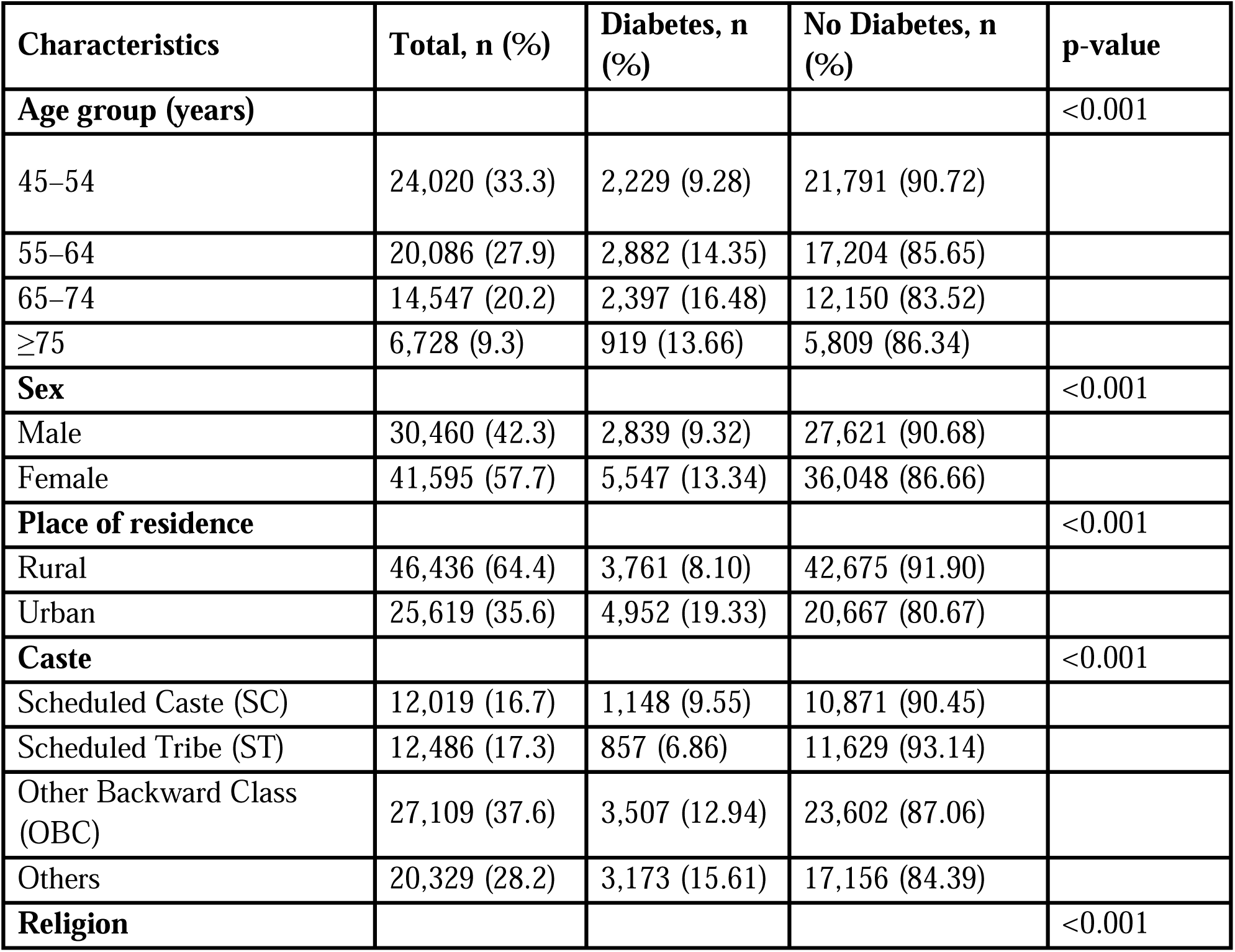
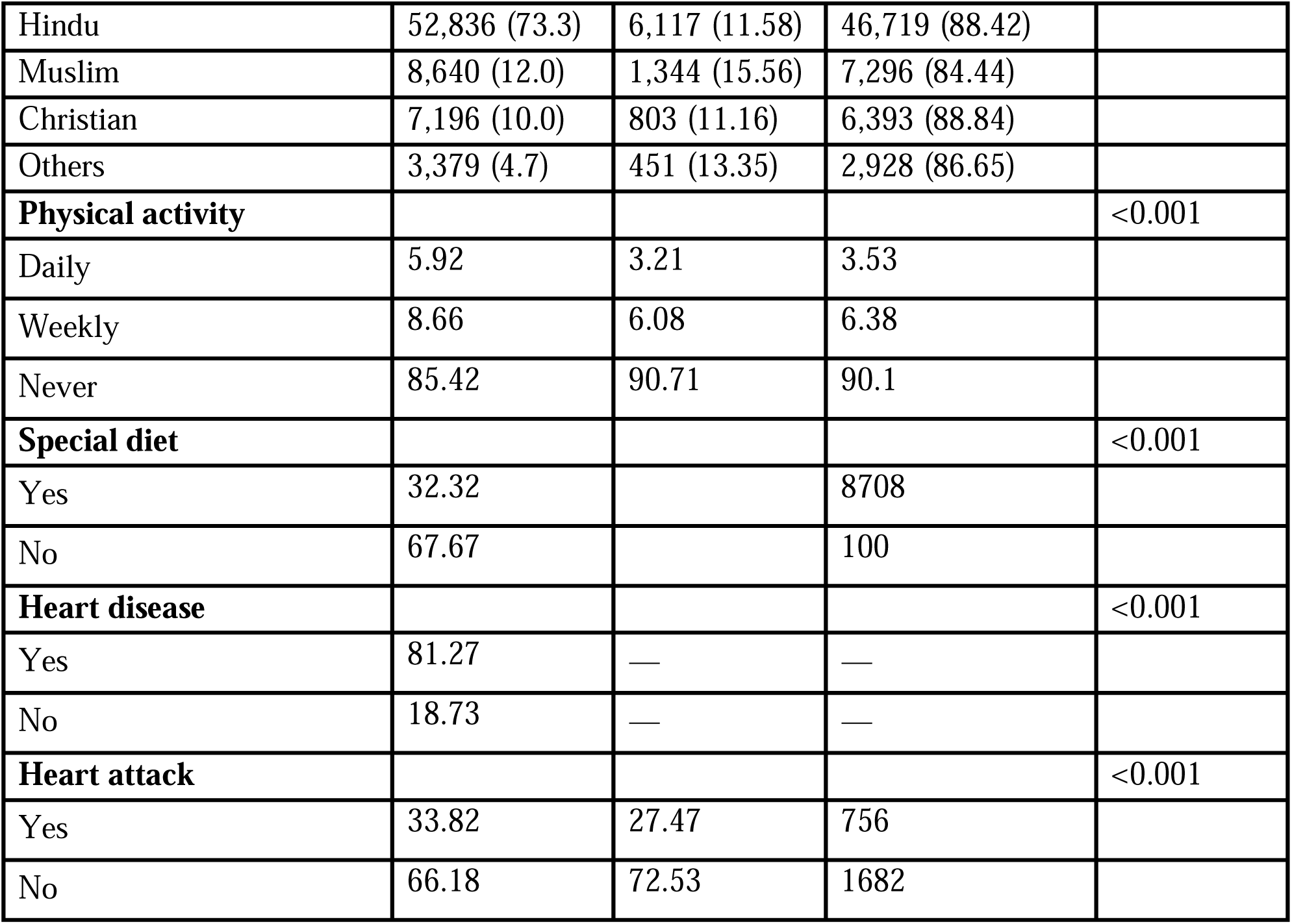
Characteristics of the Study Population and Prevalence of Diabetes among Adults Aged 45 Years and Above, India, LASI Wave-1 (2017–18)

| **Characteristics** | **Total, n (%)** | **Diabetes, n (%)** | **No Diabetes, n (%)** | **p-value** |
|---|---|---|---|---|
| **Age group (years)** |  |  |  | <0.001 |
| 45–54 | 24,020 (33.3) | 2,229 (9.28) | 21,791 (90.72) |  |
| 55–64 | 20,086 (27.9) | 2,882 (14.35) | 17,204 (85.65) |  |
| 65–74 | 14,547 (20.2) | 2,397 (16.48) | 12,150 (83.52) |  |
| ≥75 | 6,728 (9.3) | 919 (13.66) | 5,809 (86.34) |  |
| **Sex** |  |  |  | <0.001 |
| Male | 30,460 (42.3) | 2,839 (9.32) | 27,621 (90.68) |  |
| Female | 41,595 (57.7) | 5,547 (13.34) | 36,048 (86.66) |  |
| **Place of residence** |  |  |  | <0.001 |
| Rural | 46,436 (64.4) | 3,761 (8.10) | 42,675 (91.90) |  |
| Urban | 25,619 (35.6) | 4,952 (19.33) | 20,667 (80.67) |  |
| **Caste** |  |  |  | <0.001 |
| Scheduled Caste (SC) | 12,019 (16.7) | 1,148 (9.55) | 10,871 (90.45) |  |
| Scheduled Tribe (ST) | 12,486 (17.3) | 857 (6.86) | 11,629 (93.14) |  |
| Other Backward Class (OBC) | 27,109 (37.6) | 3,507 (12.94) | 23,602 (87.06) |  |
| Others | 20,329 (28.2) | 3,173 (15.61) | 17,156 (84.39) |  |
| **Religion** |  |  |  | <0.001 |
| Hindu | 52,836 (73.3) | 6,117 (11.58) | 46,719 (88.42) |  |
| Muslim | 8,640 (12.0) | 1,344 (15.56) | 7,296 (84.44) |  |
| Christian | 7,196 (10.0) | 803 (11.16) | 6,393 (88.84) |  |
| Others | 3,379 (4.7) | 451 (13.35) | 2,928 (86.65) |  |
| **Physical activity** |  |  |  | <0.001 |
| Daily | 5.92 | 3.21 | 3.53 |  |
| Weekly | 8.66 | 6.08 | 6.38 |  |
| Never | 85.42 | 90.71 | 90.1 |  |
| **Special diet** |  |  |  | <0.001 |
| Yes | 32.32 |  | 8708 |  |
| No | 67.67 |  | 100 |  |
| **Heart disease** |  |  |  | <0.001 |
| Yes | 81.27 | — | — |  |
| No | 18.73 | — | — |  |
| **Heart attack** |  |  |  | <0.001 |
| Yes | 33.82 | 27.47 | 756 |  |
| No | 66.18 | 72.53 | 1682 |  |

**Fig-3.1.**
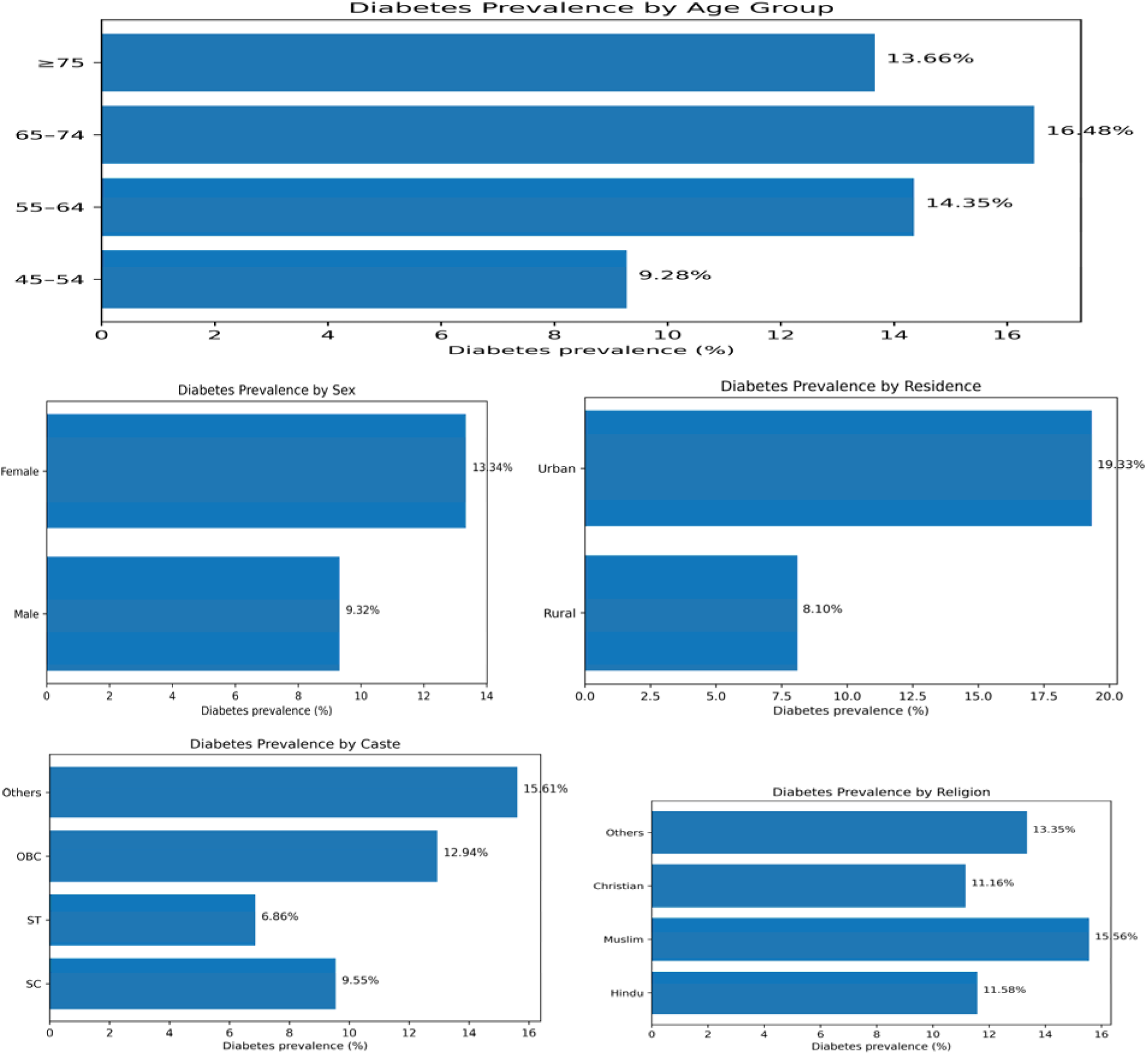
Plot for Visualization of Socio-demographic of Diabetes prevalence.

**Fig: 3.2.**
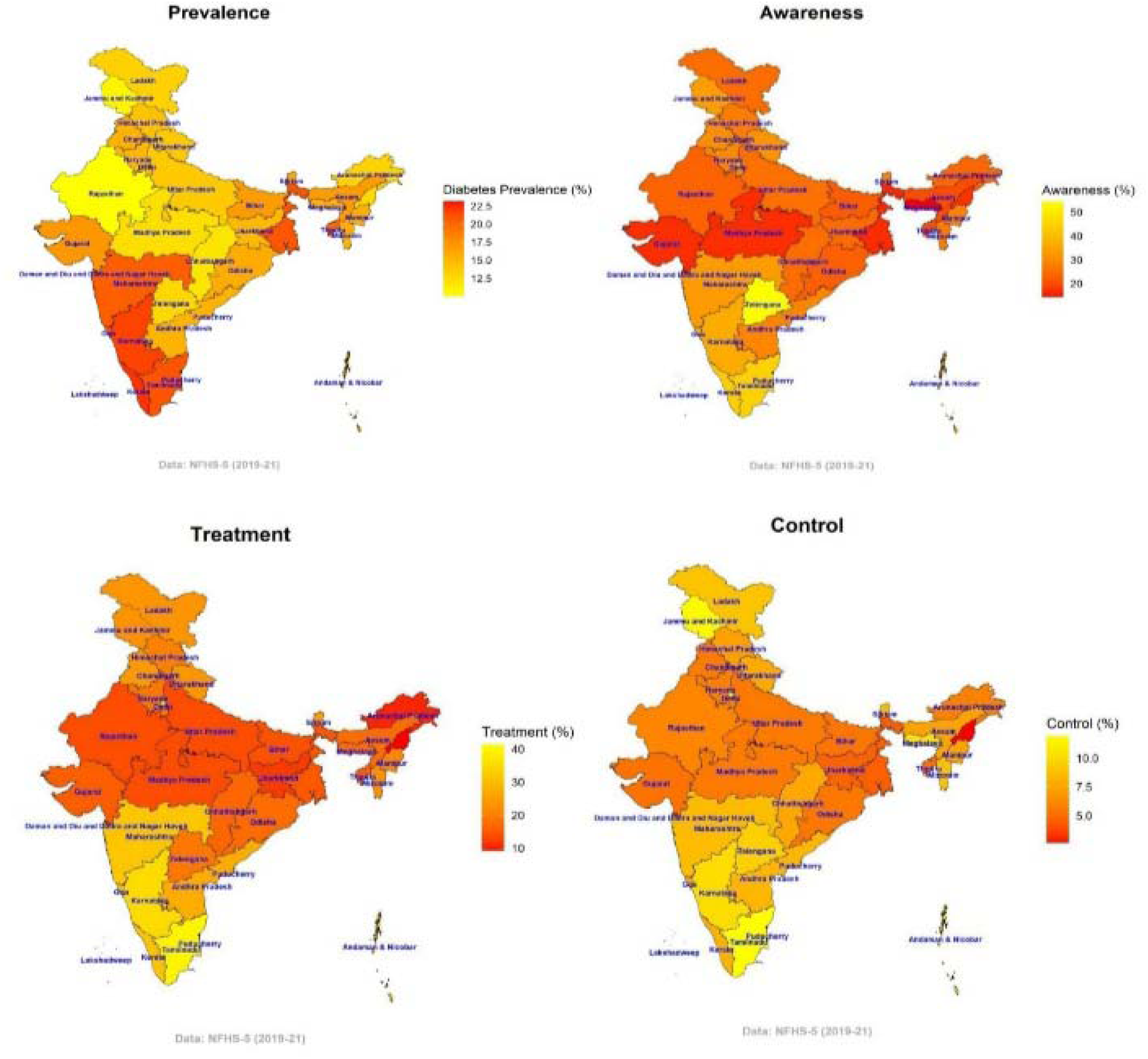
Prevalence of Diabetes in India. **Source: -**https://www.nature.com/articles/s41598-023-29978-y/figures/5

**Visualizationof Prevalence of diabetes among elderly people Based on their Physical activity/exercise, Special diet Plan as well as Heat Diseaase.**

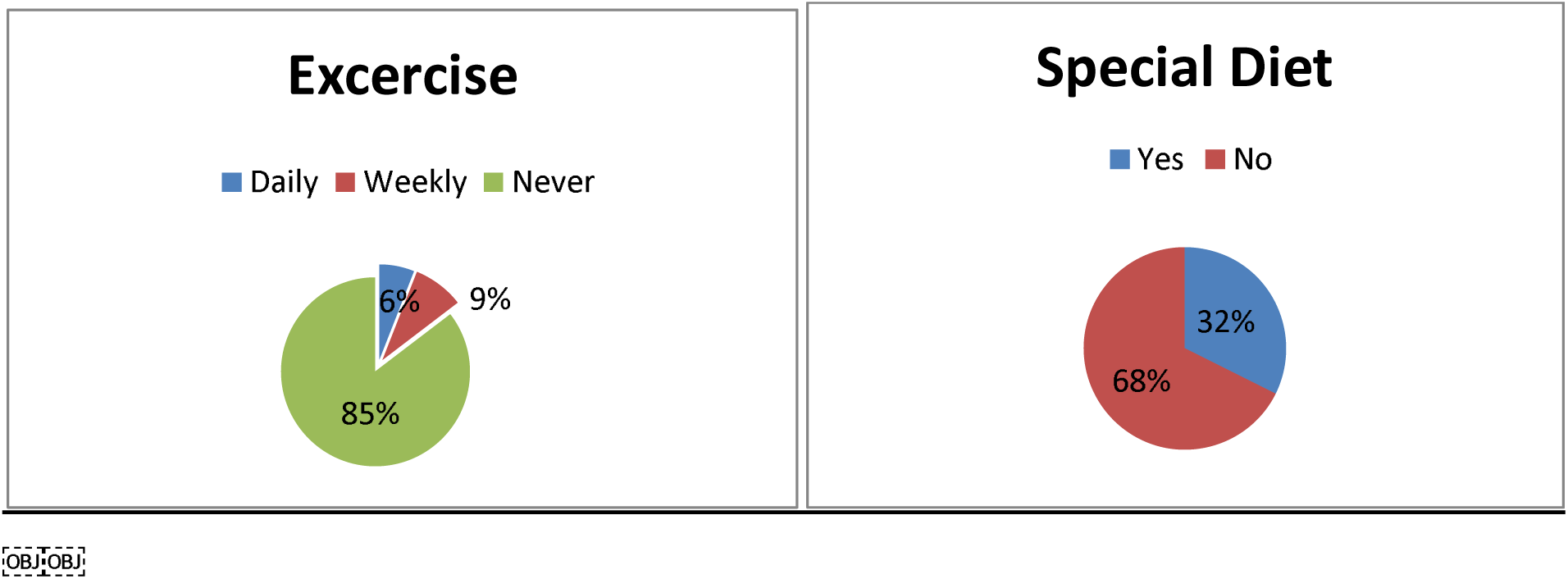

## 4 Results and Discussions

**Table 4.1.** Multivariable Logistic Regression Analysis of Factors Associated with Diabetes among Adults Aged ≥45 Years in India, LASI Wave-1 (2017–18)

| **Variable** | **Category** | **AOR** | **95% CI** | **p-value** |
|---|---|---|---|---|
| **Age group (years)** | **45–54 (Reference)** | 1.00 | — | — |
|  | 55–64 | 2.40 | 1.22–4.71 | <0.001 |
|  | 65–74 | 2.48 | 1.26–4.88 | <0.001 |
|  | ≥75 | 2.27 | 1.13–4.55 | 0.021 |
| **Sex** | **Male (Reference)** | 1.00 | — | — |
|  | Female | 1.13 | 0.94–1.35 | 0.196 |
| **Place of residence** | **Rural (Reference)** | 1.00 | — | — |
|  | Urban | 2.30 | 1.92–2.76 | <0.001 |
| **Caste** | **Scheduled Caste (Reference)** | 1.00 | — | — |
|  | Scheduled Tribe | 1.01 | 0.64–1.58 | 0.975 |
|  | Other Backward Class | 1.53 | 1.14–2.05 | 0.005 |
|  | Others | 1.21 | 0.91–1.63 | 0.182 |
| **Religion** | **Hindu (Reference)** | 1.00 | — | — |
|  | Muslim | 1.03 | 0.82–1.30 | 0.790 |
|  | Christian | 1.69 | 1.18–2.44 | 0.004 |
|  | Others | 1.09 | 0.70–1.69 | 0.709 |
| **Heart disease** | **No (Reference)** | 1.00 | — | — |
|  | Yes | 0.77 | 0.64–0.93 | 0.006 |
| **Heart attack** | **No (Reference)** | 1.00 | — | — |
|  | Yes | 0.13 | 0.06–0.28 | <0.001 |

Table 3.1 as well as fig 3.1 presents the results of the multivariable logistic regression analysis examining factors associated with diabetes among adults aged 45 years and above in India. After adjusting for potential confounding variables, **age**, **place of residence**, **caste**, **religion**, **heart disease**, and **heart attack history** remained significantly associated with diabetes. Compared with the reference group, adults aged **55–64 years** had **2.40 times higher odds** of diabetes (AOR = 2.40; 95% CI: 1.22–4.71), while those aged **65–74 years** had the highest odds (AOR = 2.48; 95% CI: 1.26–4.88). Adults aged **75 years and above** also had significantly greater odds of diabetes (AOR = 2.27; 95% CI: 1.13–4.55), indicating an increasing likelihood of diabetes with advancing age. Urban residence was independently associated with diabetes. Participants living in **urban areas** had more than **twice the odds** of diabetes compared with those living in rural areas (AOR = 2.30; 95% CI: 1.92–2.76). With respect to social characteristics, respondents belonging to the **Other Backward Class (OBC)** had significantly higher odds of diabetes than those from the Scheduled Caste category (AOR = 1.53; 95% CI: 1.14–2.05). Similarly, **Christian** participants had higher odds of diabetes than **Hindu** participants (AOR = 1.69; 95% CI: 1.18–2.44). Regarding health-related factors, respondents reporting **heart disease** had significantly different odds of diabetes (AOR = 0.77; 95% CI: 0.64–0.93), while those with a **history of heart attack** showed markedly lower adjusted odds (AOR = 0.13; 95% CI: 0.06– 0.28). These inverse associations should be interpreted cautiously because they may reflect model specification, disease management, reverse causation, or residual confounding inherent in cross-sectional analyses. No statistically significant associations were observed for **sex**, **Scheduled Tribe (ST)**, **Muslim religion**, or **other religion categories**, as their 95% confidence intervals included the null value (AOR = 1).

**Fig. 4.1.**
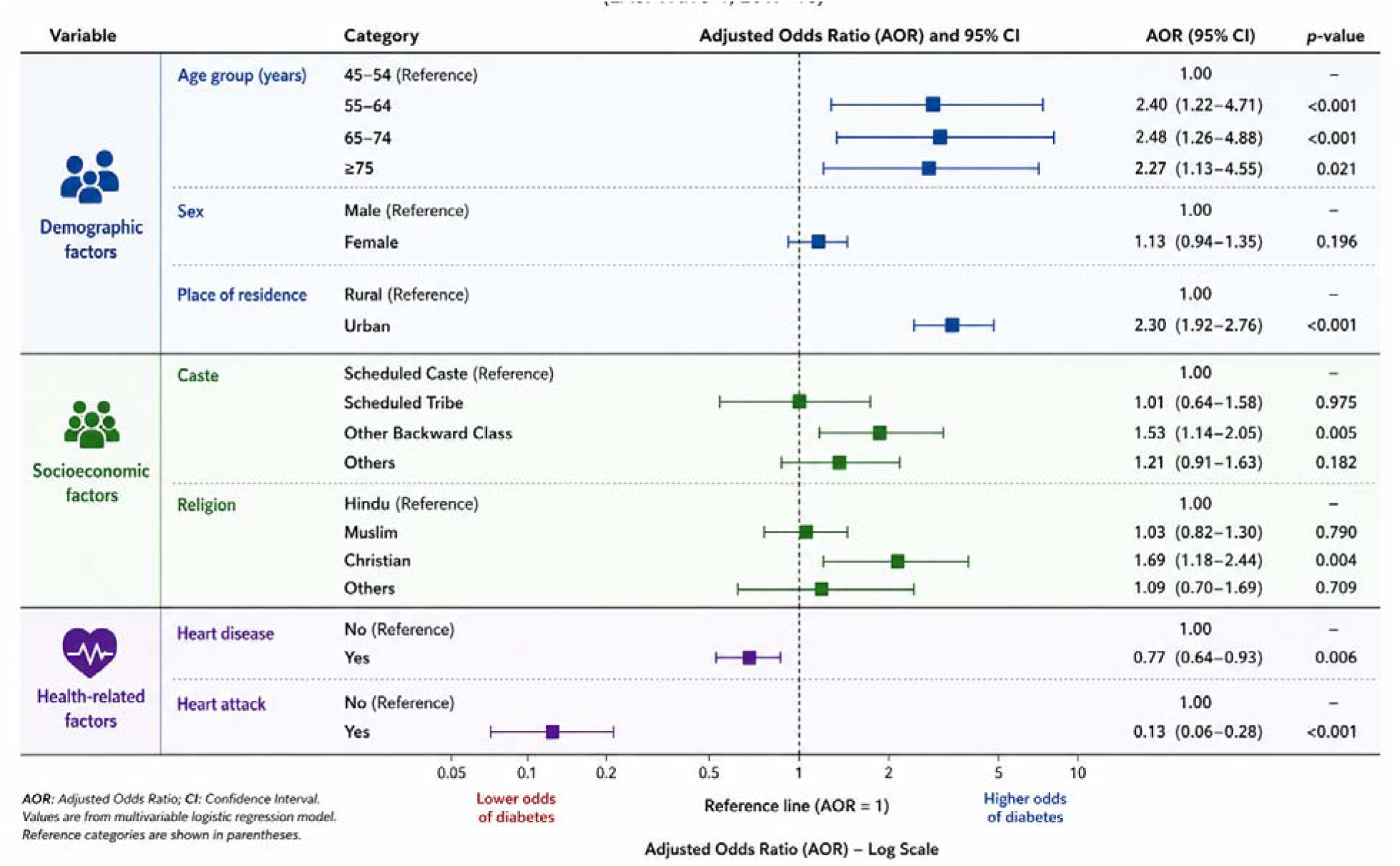
Adjusted Odds Ratio for Factor associated with diabetes among adults aged >45 Years in India.

## 4.1 Discussion

The present study examined the prevalence of diabetes and its associated demographic, socioeconomic, and health-related factors among adults aged 45 years and above in India using nationally representative data from LASI Wave-1 (2017–18). Overall, approximately one in nine participants reported having diabetes, highlighting the substantial burden of the disease among middle-aged and older adults in India.

The multivariable analysis demonstrated that increasing age was significantly associated with higher odds of diabetes. Adults aged 55 years and above had greater odds of diabetes than those aged 45–54 years, with the highest odds observed among individuals aged 65–74 years. This age-related pattern is consistent with previous Indian studies using LASI and other national datasets, which have reported that diabetes prevalence increases with age because of progressive metabolic changes, longer exposure to behavioural risk factors, and population ageing. Similar findings have also been reported in studies based on the National Family Health Survey (NFHS) and other epidemiological investigations conducted in India.

Urban residence was independently associated with higher odds of diabetes. Participants living in urban areas had more than twice the odds of diabetes compared with rural residents. This finding is consistent with earlier evidence from India, where urban populations generally experience greater exposure to sedentary lifestyles, dietary transitions, obesity, and other cardiometabolic risk factors. However, the present study cannot determine whether urban residence itself causes diabetes because the analysis is based on cross-sectional observational data.

The analysis also identified socioeconomic differences in diabetes prevalence. Participants belonging to the Other Backward Class (OBC) had significantly higher odds of diabetes than those from the Scheduled Caste category, while Christian participants showed higher odds than Hindus after adjustment for other variables. These findings may reflect differences in socioeconomic circumstances, lifestyle behaviours, healthcare utilisation, and regional variation. However, the present analysis did not directly examine these mechanisms; therefore, these associations should be interpreted cautiously.

Heart disease and previous heart attack were significantly associated with diabetes in the adjusted model. Because the outcome and covariates were measured at the same time, the temporal relationship between these conditions cannot be established. Consequently, these findings should not be interpreted as evidence that heart disease or heart attack reduces or increases the risk of diabetes. Instead, they likely reflect the complex coexistence of chronic diseases among older adults and may also be influenced by residual confounding, treatment effects, or differences in healthcare utilisation.

Contrary to the unadjusted descriptive findings, sex was not significantly associated with diabetes after adjustment for other covariates. Likewise, no statistically significant associations were observed for Scheduled Tribe (ST) or Muslim participants after controlling for potential confounders. These findings suggest that demographic and socioeconomic factors, particularly age and place of residence, explain a substantial proportion of the observed variation in diabetes prevalence.

The findings of this study should be interpreted in light of several limitations. First, the analysis is based on cross-sectional baseline data from LASI Wave-1; therefore, causal relationships cannot be inferred. Second, diabetes status was based on self-reported physician diagnosis, which may underestimate the true prevalence because undiagnosed cases were not captured. Third, information on dietary intake, medication adherence, duration of diabetes, glycaemic control, and several behavioural risk factors was not comprehensively examined in the present analysis. Despite these limitations, the study benefits from the use of a large, nationally representative sample, allowing the findings to be generalised to the Indian population aged 45 years and above. Overall, the findings identify important demographic and socioeconomic groups with a higher prevalence of diabetes and provide evidence that may assist in prioritising screening and preventive strategies among middle-aged and older adults. However, further longitudinal analyses using subsequent waves of LASI are required to examine temporal relationships and better understand the progression of diabetes over time.

## 5 Conclusion

This study examined the prevalence of diabetes and its associated factors among adults aged **45 years and above** in India using nationally representative data from the **Longitudinal Ageing Study in India (LASI) Wave-1 (2017–18)**. The multivariable logistic regression analysis showed that increasing age, urban residence, Other Backward Class (OBC) status, and Christian religion were independently associated with higher odds of diabetes after adjusting for potential confounding factors. In contrast, sex and several other sociodemographic characteristics were not significantly associated with diabetes after adjustment.

These findings should be interpreted as **associations rather than causal relationships**, as the analysis was based on cross-sectional baseline data from LASI. Furthermore, diabetes status was based on self-reported physician diagnosis, which may underestimate the true prevalence because undiagnosed cases were not captured. Consequently, the temporal relationship between diabetes and the associated factors cannot be established.

Despite these limitations, the study provides nationally representative evidence on demographic and socioeconomic disparities in diabetes among middle-aged and older adults in India. The findings may help identify population groups that could benefit from targeted diabetes screening and prevention initiatives. Future research using longitudinal follow-up data from subsequent LASI waves is needed to investigate temporal changes and better understand the causal pathways underlying diabetes among older adults in India.

## Data Availability

All data produced in the present study are available upon reasonable request to the authors

## References

Anjana, R. M., Pradeepa, R., Deepa, M., Datta, M., Sudha, V., Unnikrishnan, R., … & Mohan, V. (2011). Prevalence of diabetes and prediabetes (impaired fasting glucose and/or impaired glucose tolerance) in urban and rural India: Phase I results of the Indian Council of Medical Research–INdiaDIABetes (ICMR–INDIAB) study. Diabetologia, 54(12), 3022–3027. 10.1007/s00125-011-2291-5

Anjana, R. M. et al. Prevalence of diabetes and prediabetes in 15 states of India: Results from the ICMR–INDIAB population-based cross-sectional study. Lancet Diabetes Endocrinol. 5, 585–596 (2017).

Bjork, S., Kapur, A., King, H., Nair, J., & Ramachandran, A. (2003). Global policy: aspects of diabetes in India. Health policy, 66(1), 61–72.

Centers for Disease Control and Prevention. (2023). Diabetes meal planning. https://www.cdc.gov/diabetes/managing/eat-well/meal-plan-method.html.

Dal Canto, E., et al. Diabetes as a cardiovascular risk factor: An overview of global trends of macro and micro vascular complications. Eur. J. Prev. Cardiol. 26, 25–32 (2019).

Deshpande, A. D., Harris-Hayes, M. & Schootman, M. Epidemiology of diabetes and diabetes-related complications. Phys. Ther. 88, 1254–1264 (2008).

Federation, I. D. IDF Diabetes Atlas 10th Edition. Preprint at (2021).

Geldsetzer, P. et al. Diabetes and hypertension in India a nationally representative study of 1.3 million adults. JAMA Intern. Med. 178, 363–372 (2018).

Gyawali, B. et al. Awareness, prevalence, treatment, and control of type 2 diabetes in a semi-urban area of Nepal: Findings from a cross-sectional study conducted as a part of COBIN-D trial. PLoS ONE 13, 1 (2018).

Hu, F. B. (2011). Globalization of diabetes: the role of diet, lifestyle, and genes. Diabetes Care, 34(6), 1249–1257. 10.2337/dc11-0442

Hunt, D., Hemmingsen, B., A. M.-T. L. D. & & 2021, undefined. The WHO Global Diabetes Compact: a new initiative to support people living with diabetes. thelancet.com.

Khan, M. A. B. et al. Epidemiology of type 2 diabetes—Global burden of disease and forecasted trends. J. Epidemiol. Glob. Health 10, 107 (2020).

Krishnan, A. et al. Preparedness of primary and secondary health facilities in India to address major noncommunicable diseases: results of a National Noncommunicable Disease Monitoring Survey (NNMS). BMC Health Serv. Res. 21, 1 (2021).

Kumar, A., Gangwar, R., Ahmad Zargar, A., Kumar, R., & Sharma, A. (2024). Prevalence of diabetes in India: A review of IDF diabetes atlas 10th edition. Current diabetes reviews, 20(1), 105–114.

LASI. (2017–2018). Longitudinal Aging Study in India – Wave 1 Data. Retrieved from https://www.iipsindia.ac.in/content/lasi-wave-i

Lee, J. et al. Prevalence, awareness, treatment, and control of diabetes mellitus by depressive symptom severity: A cross-sectional analysis of NHANES 2011–2016. BMJ Open Diab. Res. Care 9, 1 (2021).

Lin, X. et al. Global, regional, and national burden and trend of diabetes in 195 countries and territories: an analysis from 1990 to 2025. Sci. Rep. 10, 1 (2020).

Liu, X. et al. Prevalence, awareness, treatment, control of type 2 diabetes mellitus and risk factors in Chinese rural population: The RuralDiab study. Sci. Rep. 6, 1 (2016).

Maiti, S., Akhtar, S., Upadhyay, A.K. et al. Socioeconomic inequality in awareness, treatment and control of diabetes among adults in India: Evidence from National Family Health Survey of India (NFHS), 2019–2021. Sci Rep 13, 2971 (2023). 10.1038/s41598-023-29978-y

Misra, A. et al. Diabetes in developing countries. J. Diabetes 11, 522–539 (2019).

Misra, A., & Khurana, L. (2008). Obesity and the metabolic syndrome in developing countries. The Journal of Clinical Endocrinology & Metabolism, 93(11_supplement_1), s9–s30. 10.1210/jc.2008-1595

Mathur, P., Leburu, S. & Kulothungan, V. Prevalence, awareness, treatment and control of diabetes in India from the countrywide national NCD monitoring survey. Front. Public Health 10, 205 (2022).

Mohan, V., Sandeep, S., Deepa, R., Shah, B., & Varghese, C. (2007). Epidemiology of type 2 diabetes: Indian scenario. Indian Journal of Medical Research, 125(3), 217–230.

Mohan, V. et al. Current status of management, control, complications and psychosocial aspects of patients with diabetes in India: Results from the DiabCare India 2011 Study. Indian J. Endocrinol. Metab. 18, 370–378 (2014).

Murugesan, N., Snehalatha, C., Shobhana, R., Roglic, G., & Ramachandran, A. (2007). Awareness about diabetes and its complications in the general and diabetic population in a city in southern India. Diabetes research and clinical practice, 77(3), 433–437.

Mutyambizi, C., Pavlova, M., Chola, L., Hongoro, C. & Groot, W. Cost of diabetes mellitus in Africa: A systematic review of existing literature. Global Health 14, 1–13 (2018).

Nanditha, A., Ma, R. C., Ramachandran, A., Snehalatha, C., Chan, J. C., Chia, K. S., … & Zimmet, P. Z. (2016). Diabetes in Asia and the Pacific: implications for the global epidemic. Diabetes care, 39(3), 472–485.

National Institute of Diabetes and Digestive and Kidney Diseases. (2023). What is diabetes?https://www.niddk.nih.gov/health-information/diabetes/overview/what-is-diabetes.

National Centre for Biotechnology Information. (2018). Ethical issues in research involving elderly people. https://www.ncbi.nlm.nih.gov/pmc/articles/PMC6220161/

Rahman, M. S. et al. Awareness, treatment, and control of diabetes in Bangladesh: A nationwide population-based study. PLoS ONE 10, 1 (2015).

Ramachandran, A., Snehalatha, C., Shetty, A. S., & Nanditha, A. (2012). Trends in prevalence of diabetes in Asian countries. World journal of diabetes, 3(6), 110.

ResearchGate. (2023). A pie chart depicting the prevalence ratio of the main types of diabetes. https://www.researchgate.net/figure/A-pie-chart-depicting-the-prevalence-ratio-of-the-main-types-of-diabetes_fig2_344478388.

Stokes, A. et al. Prevalence and unmet need for diabetes care across the care continuum in a national sample of South African adults: Evidence from the SANHANES-1, 2011– 2012. PLoS ONE 12, e0184264 (2017).

Tandon, N. et al. The increasing burden of diabetes and variations among the states of India: the Global Burden of Disease Study 1990–2016. Lancet Glob. Health 6, e1352–e1362 (2018).

Unuofin, Jeremiah Oshiomame, and Sogolo Lucky Lebelo. “Antioxidant effects and mechanisms of medicinal plants and their bioactive compounds for the prevention and treatment of type 2 diabetes: an updated review.” Oxidative medicine and cellular longevity 2020 (2020).

Wang, Q. et al. Prevalence, awareness, treatment and control of diabetes mellitus among middle-aged and elderly people in a rural Chinese population: A cross-sectional study. PLoS ONE 13, 1 (2018).

Wong, E. et al. Diabetes and risk of physical disability in adults: a systematic review and meta-analysis. Lancet Diabetes Endocrinol. 1, 106–114 (2013).

World Health Organization. (2023). Diabetes. https://www.who.int/news-room/fact-sheets/detail/diabetes

Yoon, K. H., Lee, J. H., Kim, J. W., Cho, J. H., Choi, Y. H., Ko, S. H., … & Son, H. Y. (2006). Epidemic obesity and type 2 diabetes in Asia. The Lancet, 368(9548), 1681–1688. 10.1016/S0140-6736(06)69703-1

